# Community-level face mask usage in Boston, MA

**DOI:** 10.1101/2021.05.07.21256840

**Authors:** Meera McLane, Sharon M. Casey, Allison Gallagher, Maame Akosua Ohemeng-Tinyase, Sabina Yosif, Sree Ramya Chaparala, Sarah Lincoln, Eleanor J. Murray

## Abstract

**What is already known about this topic?:** Community-level face mask use is encouraged as an important preventive measure against COVID-19 transmission, and evidence suggests that jurisdictions which implement face mask mandates see a subsequent decline in COVID incidence.

**What is added by this report?:** In the Greater Boston area when a face mask mandate is in effect, 95% of people observed were wearing some type of face covering. Most of which were wearing fabric/cloth coverings (51%) or single use surgical masks (40%). Of those wearing a face covering, 85% were appropriately fitted. Indoor locations have higher adherence of appropriately worn face masks, compared to outdoor locations.

**What are the implications for public health practice?:** Adherence with face mask mandates was very high, but many individuals wore fabric face masks with unknown filtration efficacy. In addition, it was common for individuals to mis-wear, adjust, or remove their masks. Public health policies requiring mask use should include messaging about appropriate type and best practices for use.

## Introduction

Early strategies to reduce the spread of SARS-CoV-2 focused on non-pharmaceutical interventions, including personal protective equipment (PPE) [1,2].While evidence supports the use of face coverings in high-risk settings [3], data from community-level settings are limited. Recent data suggests that jurisdictions with mask mandates experienced a decrease in COVID-19 incidence [3]. The high transmissibility of SARS-CoV-2, combined with asymptomatic and pre-symptomatic transmission, strengthens the recommendation for universal use of face coverings [4,5], but an ongoing shortage of medical-grade masks has led to advocacy for cloth face coverings among the general public.

Several challenges are associated with the use of fabric face coverings. First, due to unknown design and filtration capacity, the efficacy of cloth masks is unclear [6]. Second, there has been a lack of guidance on appropriate mask wearing techniques. Third, concerns exist about risk compensation behavior associated with mask use. This study observed the use and adherence of masks among communities in the Greater Boston area of Massachusetts. Ninety-five percent of people observed were wearing masks and 85% wore them appropriately. Interestingly, our results show a high level of agreement between levels and appropriateness of mask wearing in the Boston area with those reported for Louisville Kentucky [7]. Results of our study will contribute to understanding the heterogeneity and adherence of mask wearing in the community and how this might reduce COVID-19 transmission.

## Methods

Data were collected by in-person observations between October and December 2020 at both outdoor and indoor locations with safe areas for observation, including public areas, grocery stores, an airport, and public transportation (buses and trains). Six study personnel recorded data from adults and adolescents of perceived mask-wearing age according to CDC guidelines, either on paper or electronically, on varying days and times, including mornings, afternoons, and evenings, on weekdays and weekends. Each observer recorded data at a single location for 10 minutes per session. Study personnel recorded face covering data for individuals they could directly see. Observers remained stationary in locations, where possible, however several indoor locations required the observer to move to maintain safe distance. Individuals were recorded as one observation per session. Each observer pre-specified observation rules at the start of the observation session, which were location and setting specific. Some examples include “record every individual in the enclosed space” or “individuals that enter this space during the session”. For indoor sessions, individuals observed inside or walking outside directly towards the observer were included. Decision factors also depended on how busy the area being recorded was.

Data recorded included 1) if an individual was wearing any face covering (yes/no), 2) type of covering (N-95-type; surgical type; fabric/cloth; neck buff/gaiter; t-shirt/scarf; or other), 3) number of face coverings worn, 4) how the covering was worn/fitted (appropriately - according to CDC guidelines, completely covered the nose and mouth and was secured under the chin; appropriate but loose - may not cover nose appropriately; below the nose; below the mouth/on chin; or other), 5) touching of the face covering (pulling on/off). Other PPE, including face-shields and goggles, was also recorded. Data on children perceived by the study member to be less than 2 years were not collected as they were exempt from these local guidelines. No data on perceived gender identity/sex, race/ethnicity, or age were collected in an effort to avoid stigmatization of any sub-populations, and due to challenges with observer-perception of these characteristics. The Institutional Review Board at Boston University determined this study as non-human subject research. Data were compiled into a Google Sheet document and verified for accuracy. Analyses were conducted using SAS (version 9.4; SAS Studio) and figures were generated using GraphPad Prism version 8.4 (GraphPad Software Inc, San Diego, CA).

## Results

Observers conducted 40 10-minute sessions at 12 sites around the Greater Boston area (Fig 1), during which 1,517 observations were recorded. Ninety-five percent of people observed were wearing some type of face covering (n=1,447; Fig 2A). Fabric/cloth coverings (51%) and single use surgical masks (40%) were the most prevalent, followed by N95-type masks (4%), neck buff type (4%), or other face coverings such as bandanas (Table 1). Twelve people were recorded as wearing more than one covering, 2 with goggles, and 1 with a face-shield.

**Figure 1.**
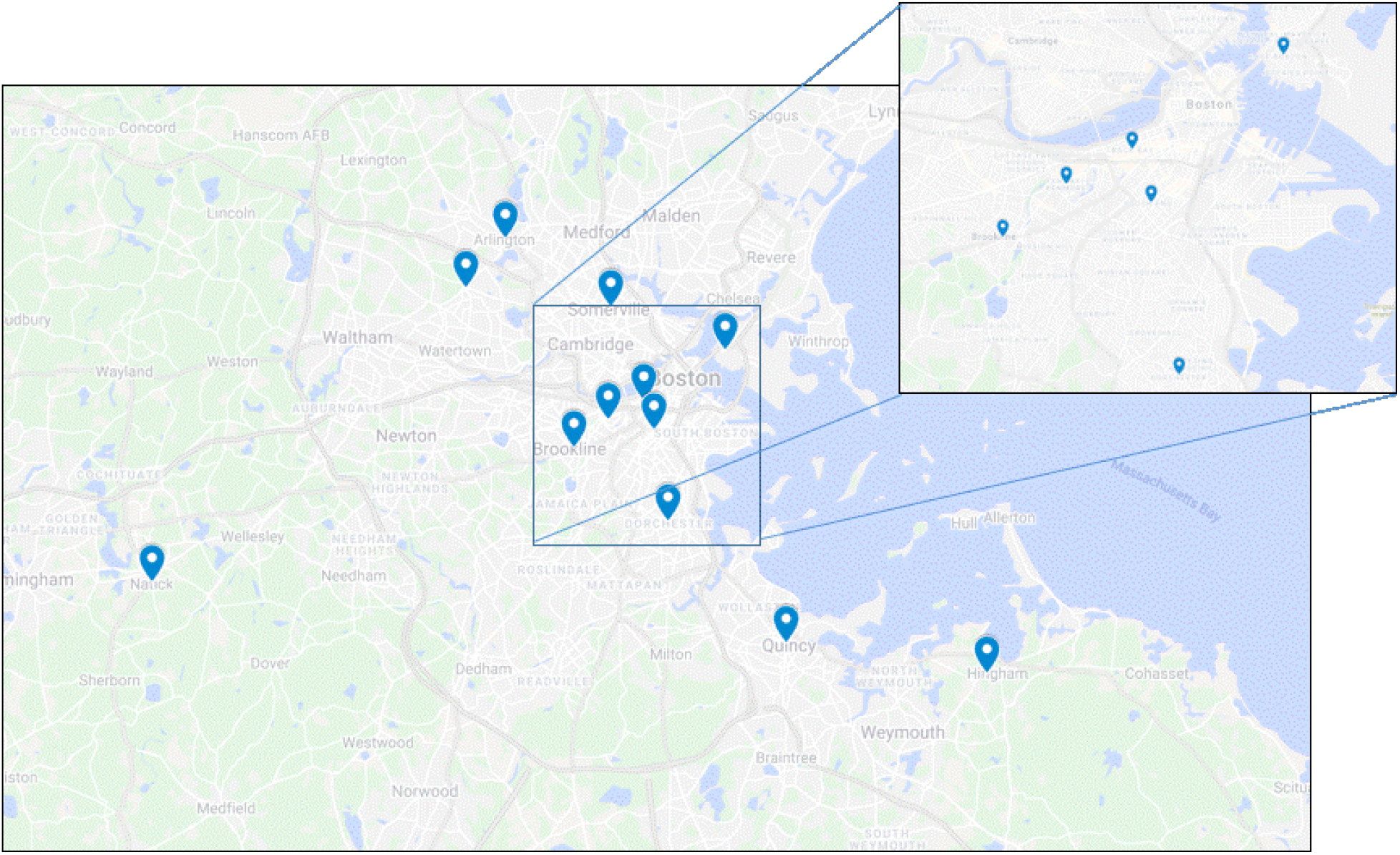
Map of data collection sites, Greater Boston area, Massachusetts.

**Figure 2B.**
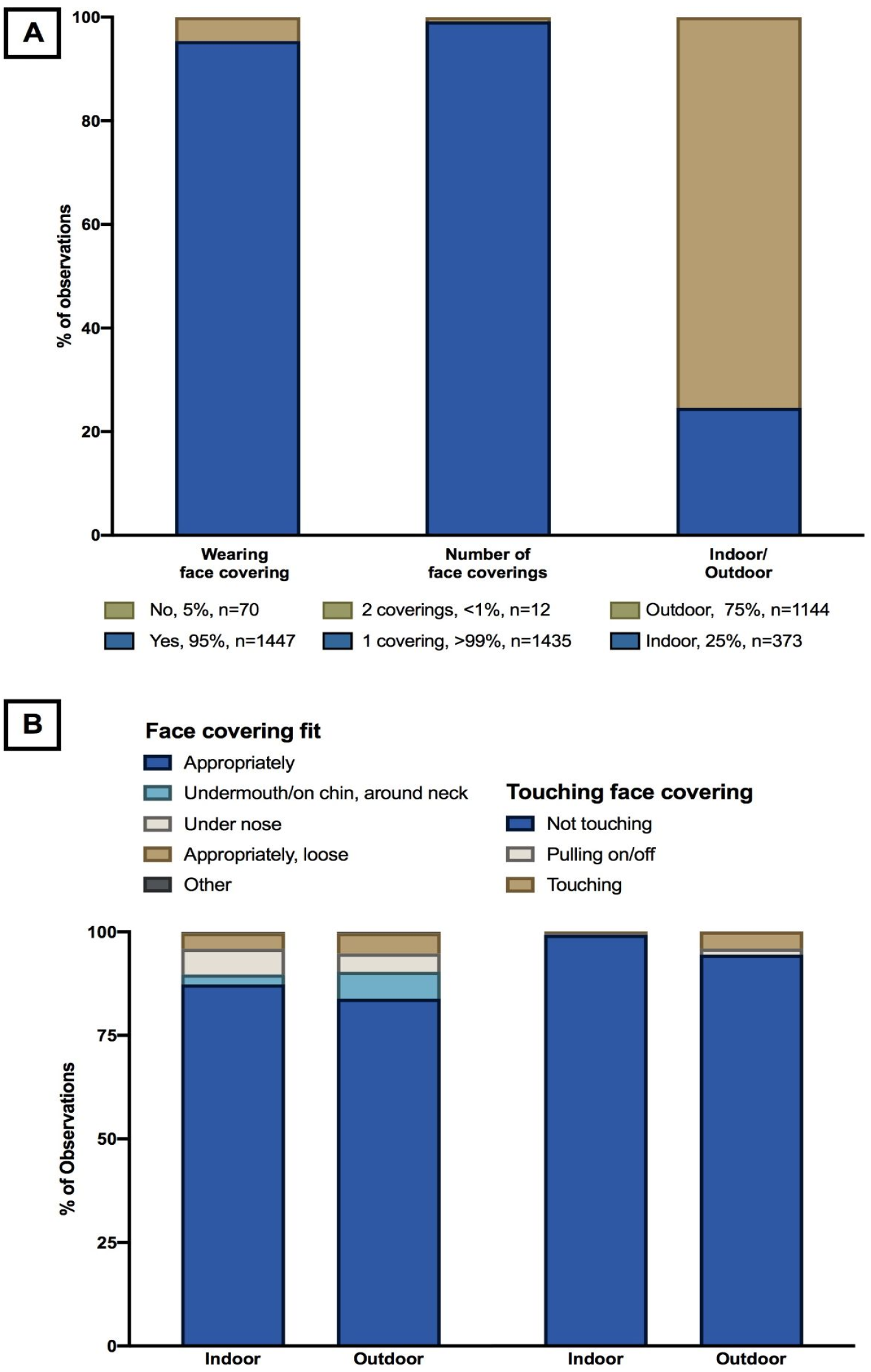
Face mask adherence. (A) Percentage of observations wearing a face covering, number of face coverings, and indoor/outdoor observation sessions. (B) Face covering fit and touching, stratified by indoor versus outdoor location.

**Table 1.** Frequency of Face Covering Type.

| Face Covering Type | (n=1,447/1,517, %) |
| --- | --- |
| N95/KN95-type | 56 (3.9) |
| Surgical | 572 (39.9) |
| Fabric or cloth | 743 (51.7) |
| Neck buff | 54 (3.7) |
| T-shirt or scarf | 6 (0.4) |
| Other face covering* | 4 (0.3) |

\*Bandana

Appropriately worn face masks were defined and recorded as fitted close to the face, covering both the mouth and nose. In this sample, 85% wore appropriately fitted coverings; the remaining 15% wore the covering under their nose, too loosely, or below their mouth (Fig 2B). Most were not touching their face covering (94%). Slightly more people were touching the face covering and/or wore it inappropriately when outdoors versus indoors (Fig 2B).

## Discussion

Evidence collected during the COVID-19 pandemic suggests that community mask wearing is an effective strategy for controlling infectious diseases, including COVID-19 (4,6). However, most research on the impacts of face mask use neglects to consider heterogeneity in mask use, type, or method. This study collected these data in local settings to improve the ability to model the impact of face mask policies on COVID-19 and other respiratory diseases.

Face mask usage was very high in the Boston area during the study period. However, the most common type was fabric masks. The high proportion of individuals wearing these types of face coverings highlights the importance of accounting for varying efficacy when studying the impact of mask use. In addition, although mask use was generally good, many individuals were observed to be wearing ill-fitting masks, wearing masks inappropriately, or adjusting or removing their masks during our observation periods. This suggests a need for improved messaging about the appropriate method for wearing, doffing, and donning face masks to ensure maximum effectiveness, as well as the importance of considering imperfect use when modeling the impact of mask mandates and other mask interventions.

Our data are an important contribution to the understanding of and practices related to mask wearing. We encouraged that other researchers conduct similar surveys of their local populations. Our survey tool can be found online at https://osf.io/7dy54/ to facilitate other data collection efforts. The dataset used in this study can be shared with researchers who have the means to conduct larger-scale investigations of face mask wearing.

Face covering use is likely to vary widely between geographical regions and political jurisdictions, and it is anticipated that the results are not widely generalizable outside the Boston, MA area. However, our findings show a high degree of similarity between mask wearing practices with previously reported mask wearing practices in indoor settings in Louisville Kentucky [7]. Additional data on other jurisdictions is needed to understand how mask wearing practices may change over the course of an outbreak, and how they vary by local political environments.

## Data Availability

Our survey tool can be found online at https://osf.io/7dy54/ to facilitate other data collection efforts. The dataset used in this study can be shared with researchers who have the means to conduct larger-scale investigations of face mask wearing.

## References

1. CDC. Coronavirus Disease 2019 (COVID-19). Centers for Disease Control and Prevention. Published February 11, 2020. Accessed April 30, 2021. https://www.cdc.gov/coronavirus/2019-ncov/science/science-briefs/masking-science-sars-cov2.html

2. Honein MA. Summary of Guidance for Public Health Strategies to Address High Levels of Community Transmission of SARS-CoV-2 and Related Deaths, December 2020. MMWR Morb Mortal Wkly Rep. 2020;69. doi:10.15585/mmwr.mm6949e2

3. Fischer CB, Adrien N, Silguero JJ, Hopper JJ, Chowdhury AI, Werler MM. Mask adherence and rate of COVID-19 across the United States. PLOS ONE. 2021;16(4):e0249891. doi:10.1371/journal.pone.0249891

4. Lerner AM, Folkers GK, Fauci AS. Preventing the Spread of SARS-CoV-2 With Masks and Other “Low-tech” Interventions. JAMA. 2020;324(19):1935. doi:10.1001/jama.2020.21946

5. Wang J, Pan L, Tang S, Ji JS, Shi X. Mask use during COVID-19: A risk adjusted strategy. Environ Pollut Barking Essex 1987. 2020;266:115099. doi:10.1016/j.envpol.2020.115099

6. Jefferson T, Jones M, Al-Ansary L, et al. Physical Interventions to Interrupt or Reduce the Spread of Respiratory Viruses. Part 1 - Face Masks, Eye Protection and Person Distancing: Systematic Review and Meta-Analysis. Public and Global Health; 2020. doi:10.1101/2020.03.30.20047217

7. Karimi, S, Salunkhe S, White K, et al. Prevalence of Unmasked and Improperly Masked Behavior in Indoor Public Areas during the COVID-19 Pandemic: Analysis of a Stratified Random Sample from Louisville, Kentucky. PLOS ONE Rev. Published online 2021.

